# Machine Learning Algorithms for Neurosurgical Preoperative Planning: A Comprehensive Scoping Review of the Literature

**DOI:** 10.1101/2024.10.04.24314930

**Authors:** Jhon E. Bocanegra-Becerra, Julia Sader Neves Ferreira, Gabriel Simoni, Anthony Hong, Wagner Rios-Garcia, Mohammad Mirahmadi Eraghi, Adriam M. Castilla-Encinas, Jhair Alejandro Colan, Rolando Rojas-Apaza, Emanuel Eduardo Franco Pariasca Trevejo, Raphael Bertani, Miguel Angel Lopez-Gonzalez

**Affiliations:** Academic Department of Surgery, School of Medicine, Universidad Peruana Cayetano Heredia, Lima, Peru; Santo Amaro University, Brazil; Centro Universitário de Várzea Grande, Várzea Grande, MT, Brasil; University of Costa Rica, Costa Rica; Facultad de Medicina Humana, Universidad Nacional San Luis Gonzaga, Ica, Peru; Student Research Committee, School of Medicine, Islamic Azad University, Qeshm International Branch, Qeshm, Iran; Sociedad Científica de San Fernando, Universidad Nacional Mayor de San Marcos, Lima, Peru; School of Medicine, University of Pittsburgh, PA, USA; Department of Neurosurgery, Hospital Edgardo Rebagliati Martins. Essalud, Lima, Peru; Grupo de Inteligencia Artificial PUCP-IA-PUCP, Pontificia Universidad Católica del Peru (PUCP), Lima, Perú; Department of Neurosurgery, University of Sao Paulo, Brazil; Department of Neurosurgery, Loma Linda University Medical Center, Loma Linda, CA, USA

**Keywords:** Machine Learning, Neurosurgery, Spine Surgery, Artificial Intelligence

## Abstract

**Introduction:** Preoperative neurosurgical planning is a keen step to avoiding surgical complications, reducing morbidity, and improving patient safety. The incursion of machine learning (ML) in this domain has recently gained attention, given the notable advantages in processing large data sets and potentially generating efficient and accurate algorithms in patient care.

**Objective:** To evaluate the evolving applications of ML algorithms in the preoperative planning of brain and spine surgery.

**Methods:** In accordance with the Arksey and O’Malley framework, a scoping review was conducted using three databases (Pubmed, Embase, and Web of Science). Articles that described the use of ML for preoperative planning in brain and spine surgery were included. Relevant data were collected regarding the neurosurgical field of application, patient baseline features, disease description, type of ML technology, study’s aim, preoperative ML algorithm description, and advantages and limitations of ML algorithms.

**Results:** Our search strategy yielded 7,407 articles, of which 8 studies (5 retrospective, 2 prospective, and 1 experimental study) satisfied the inclusion criteria. Clinical information from 518 patients (62.7% female; mean age: 44.8 years) was used for generating ML algorithms, including convolutional neural network (14.3%), logistic regression (14.3%), random forest (14.3%), and other algorithms (Table 1). Neurosurgical fields of applications included functional neurosurgery (37.5%), tumor surgery (37.5%), and spine surgery (25%). The main advantages of ML included automated processing of clinical and imaging information, selection of an individualized patient surgical approach and data-driven support for treatment decision-making. All studies reported technical limitations, such as long processing time, algorithmic bias, limited generalizability, and the need for database updating and maintenance.

**Conclusion:** ML algorithms for preoperative neurosurgical planning are being developed for efficient, automated, and safe treatment decision-making. Enhancing the robustness, transparency, and understanding of ML applications will be crucial for their successful integration into neurosurgical practice.

## Introduction

Preoperative neurosurgical planning helps avoid surgical complications, reduce morbidity, and increase patient safety.^1–3^ In addition, this surgical stage allows neurosurgeons to conceive contingency strategies.^4,5^ However, neurosurgical planning may require accurate analysis of multiple sources of information, such as diagnostic and functional study images.^6,7^ In this regard, machine learning (ML) algorithms offer the opportunity to process the required data efficiently for accurate and more personalized planning for each patient.^8,9^ They also produce correlations and patterns, and these results are used to predict future events.^10,11^ These techniques are used in tumor segmentation, epilepsy treatment, risk assessment, and ante position to surgical complications, among others.^12,13^

Despite promising advances, integrating ML into neurosurgical practice faces several challenges.^14–16^ These include the need for extensive, quality data, algorithm development, and the need for rigorous regulatory validation and approval.^17,18^ In addition, the practical implementation of these algorithms in clinical settings requires ethics and consideration of workflow integration, user training, and patient safety.^19–21^

While this artificial system is on the rise, addressing the aforementioned challenges will be essential for the continued evolution of ML applications in neurosurgical preoperative planning.^22^ Therefore, we aimed to conduct a scoping review to explore and evaluate the evolving applications of ML algorithms in brain and spine preoperative planning, highlighting their applications, limitations, and prospects.

## Material and methods

### Search Strategy and Study Selection

In accordance with the Arksey and O’Malley framework and the Preferred Reporting Items for Systematic Reviews and Meta-Analyses (PRISMA) extension for Scoping Reviews,^23^ three databases (PubMed, EMBASE, and SCOPUS) were queried from the date of their inception until February 2024. A PROSPERO registration was procured (registration number: CRD42024510340)

The search strategy comprised the following MESH terms, keywords, and Boolean operators: (“machine learning” OR “ML” OR “artificial intelligence” OR “AI”) AND (“planning” OR “planning” OR “pre-operative” OR “pre operative) and (“neurosurg*” OR “spine*” OR “spinal*). Furthermore, all articles’ Reference lists were also screened for additional studies and enhance the comprehensiveness of this study.

Two independent reviewers screened search results by title, abstract, and full text (W.R.G, A.M.C.E). Any discrepancies were resolved by consensus or consultation with a third reviewer (J.E.B.B). For the final report and statement of this scoping review, we utilized the PRISMA Checklist.

### Inclusion and Exclusion Criteria

We included original studies published in English that described the implementation of diverse ML algorithms in the preoperative planning of brain or spine surgeries. We excluded editorials, letters, commentaries, opinion pieces, conference abstracts, literature reviews, and articles incorporating ML into distinct categories different from pre-operative planning, such as diagnosis, risk factor prediction, or prognosis.

### Data Extraction and Statistical Analysis

Three independent reviewers (J.E.B.B., W.R.G, A.) extracted the data in a standardized collection form. Data fields included a neurosurgical field of application, patient baseline features, disease description, type of ML technology, study’s aim, preoperative ML algorithm description, and advantages and limitations of ML algorithms. The collected variables of interest were summarized as counts and proportions. Information was stored using Microsoft Excel® 2016, and descriptive statistics were performed using IBM SPSS version 29.

### Expert Consultation

Quality assessment and evaluation of good practices in ML were thoroughly assessed by a ML engineer and data scientist from the Grupo de Inteligencia Artificial PUCP-IA-PUCP, Pontificia Universidad Católica del Peru (PUCP), Lima, Peru.

## Results

Our search strategy yielded 7,407 articles, from which 1820 duplicate records were removed, and 5587 records underwent title and abstract screening. From these, 5563 were excluded, leaving 24 studies that were sought for retrieval, from which 2 reports were not retrieved, leaving 22 studies that underwent full-text assessment for eligibility. Ultimately, 8 studies (5 retrospective, 2 prospective, and 1 experimental study) met the inclusion criteria. The PRISMA flow chart that depicts the rigorous selection process is presented in **Figure 1**.

**Figure 1.**
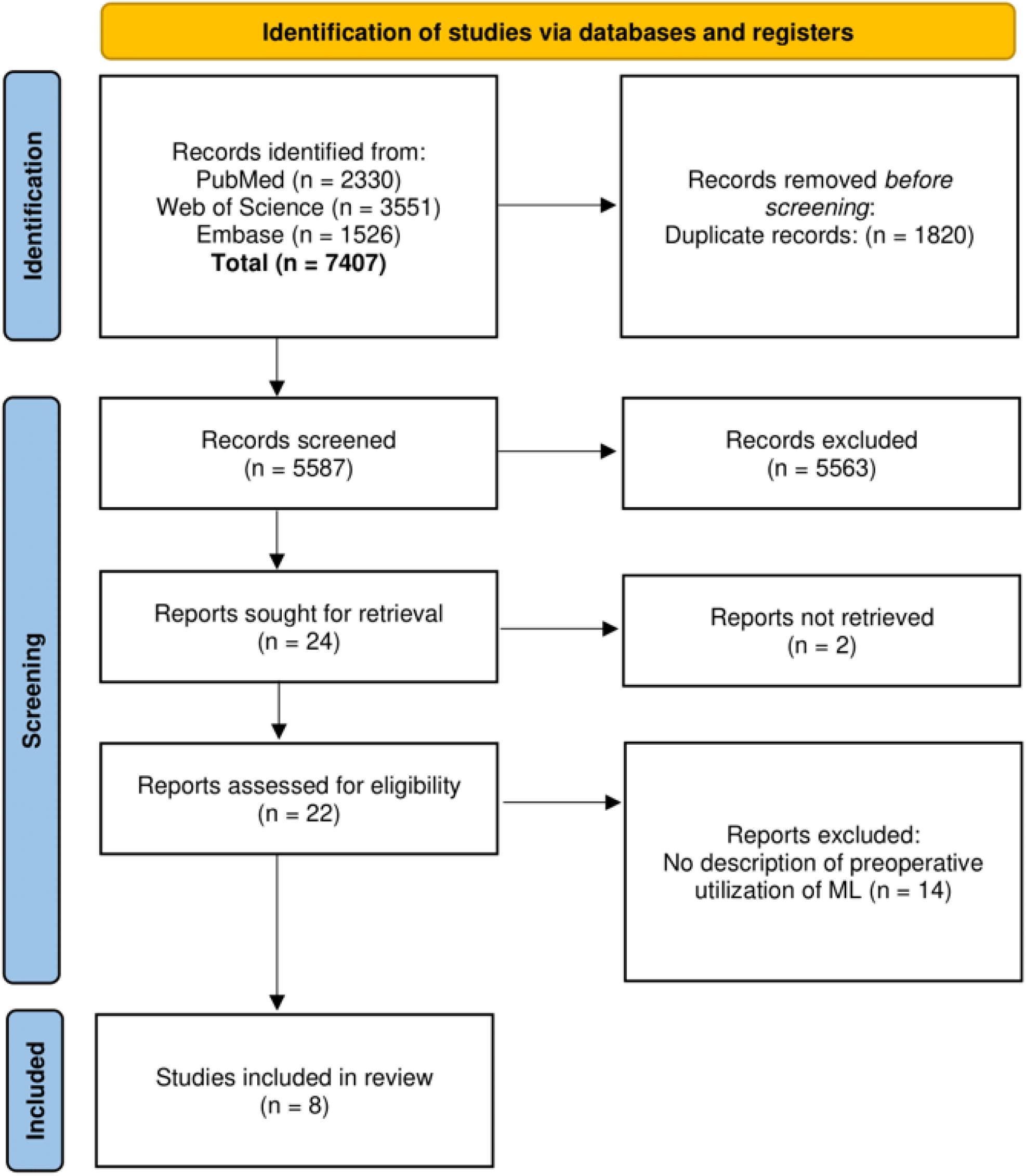
PRISMA flow diagram. ML: machine learning

The characteristics of the included studies are summarized in **Table 1**. From these, individual studies were conducted in the USA, France, Turkey, Israel, and China, and 3 studies were from Germany. Neurosurgical fields of applications included functional neurosurgery (37.5%), tumor surgery (37.5%), and spine surgery (25%) **(Figure 2)**. Clinical information from 518 patients (62.7% female; mean age: 44.8 years) was used for generating ML algorithms, including convolutional neural network (14.3%), logistic regression (14.3%), random forest (14.3%), and other algorithms (**Tables 2-3**). Based on the detailed information presented in Tables 2-3, the main advantages of ML have been summarized in three categories, including automated processing of clinical and imaging information, selection of an individualized patient surgical approach, and data-driven support for treatment decision-making **(Figure 3)**. Technical difficulties were reported by all studies, such as processing time, algorithmic bias, limited generalizability, and the need for database updating and maintenance.

**Table 1.**
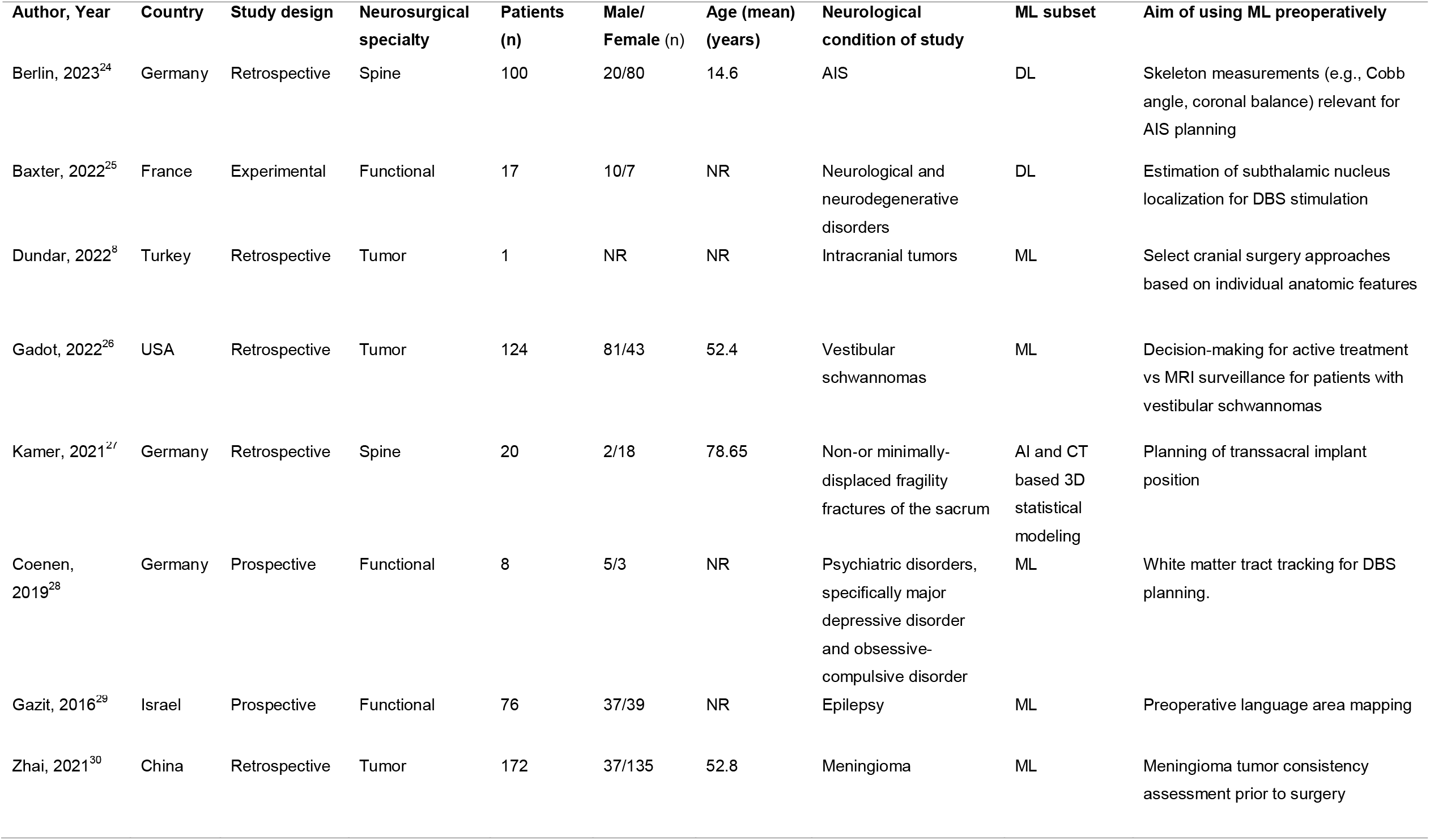
Characteristics of included studies, demographics, and clinical information. AI: artificial intelligence; AIS: adolescent idiopathic scoliosis; CT: computed tomography; DBS: deep brain stimulation; DL: deep learning; ML: machine learning; MRI: magnetic resonance imaging; PC: principal components; VS: vestibular schwannoma; 3D: Tridimensional.

**Table 2.**
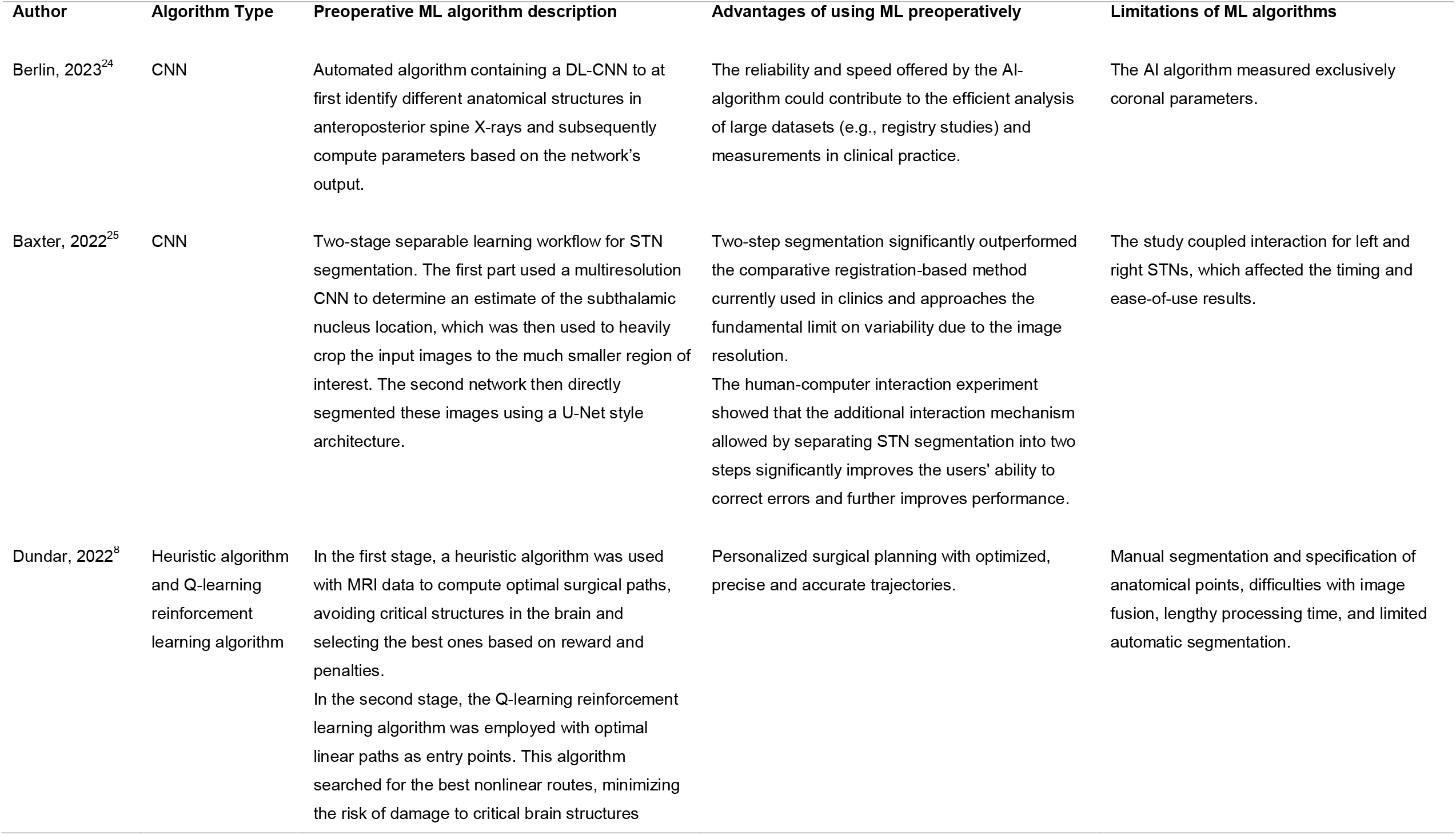
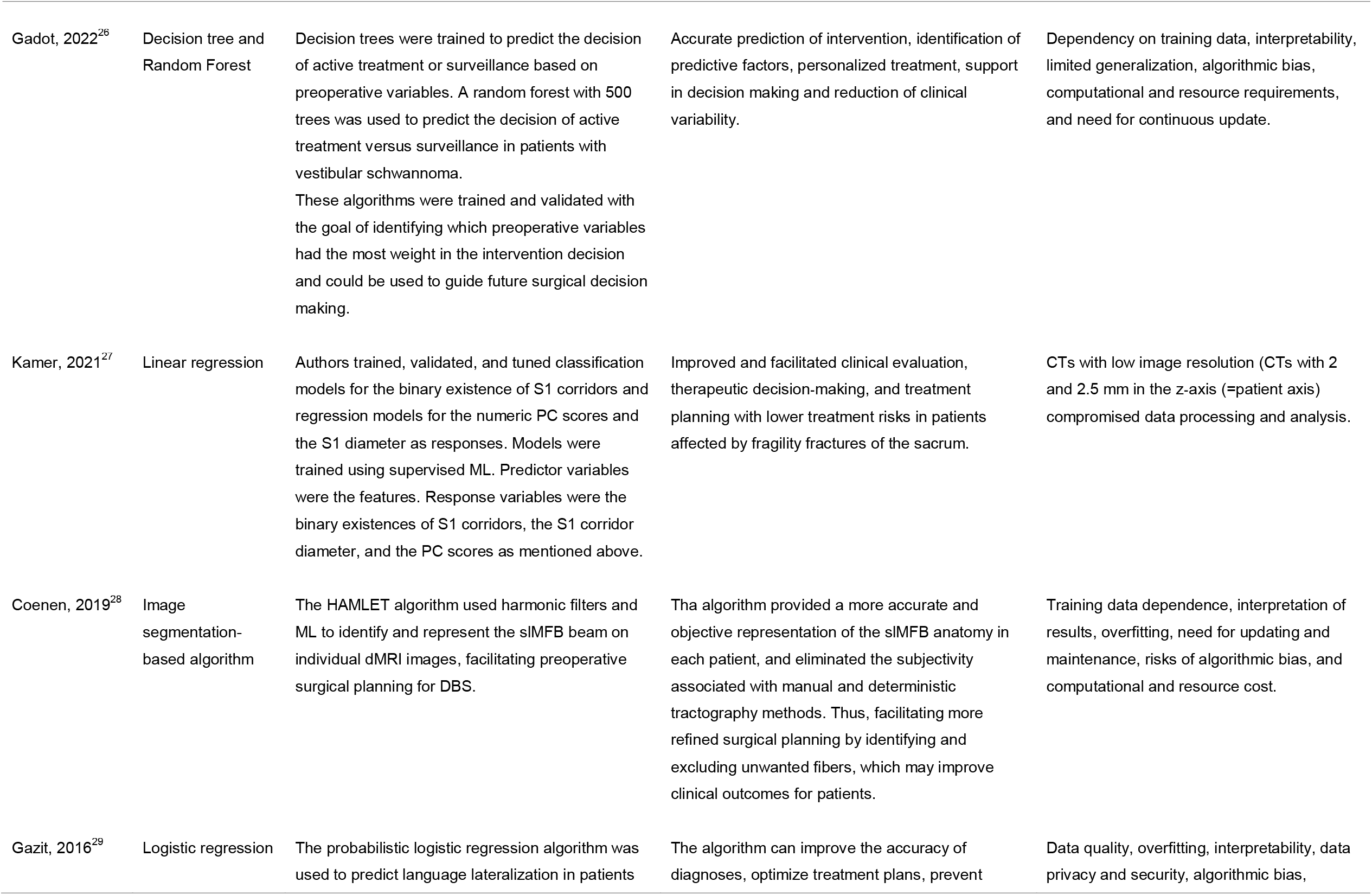
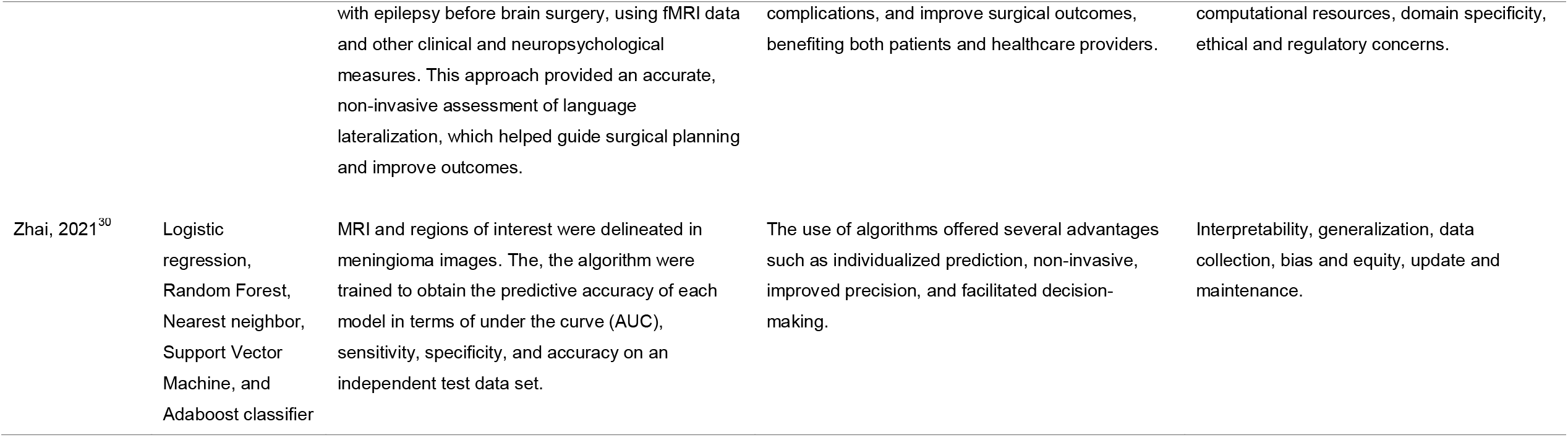
Details related to ML algorithms, advantages and limitations. AI: artificial intelligence; CNN: convoluted neural networks; CT: computed tomography; DBS: deep brain stimulation; DL: deep learning; dMRI: Diffusion magnetic resonance imaging; ML: machine learning; HAMLET: Hierarchical Harmonic Filters for Learning Tracts from Diffusion MRI; MRI: magnetic resonance imaging; STN: subthalamic nucleus; slMFB: superolateral medial forebrain bundle

**Table 3.**
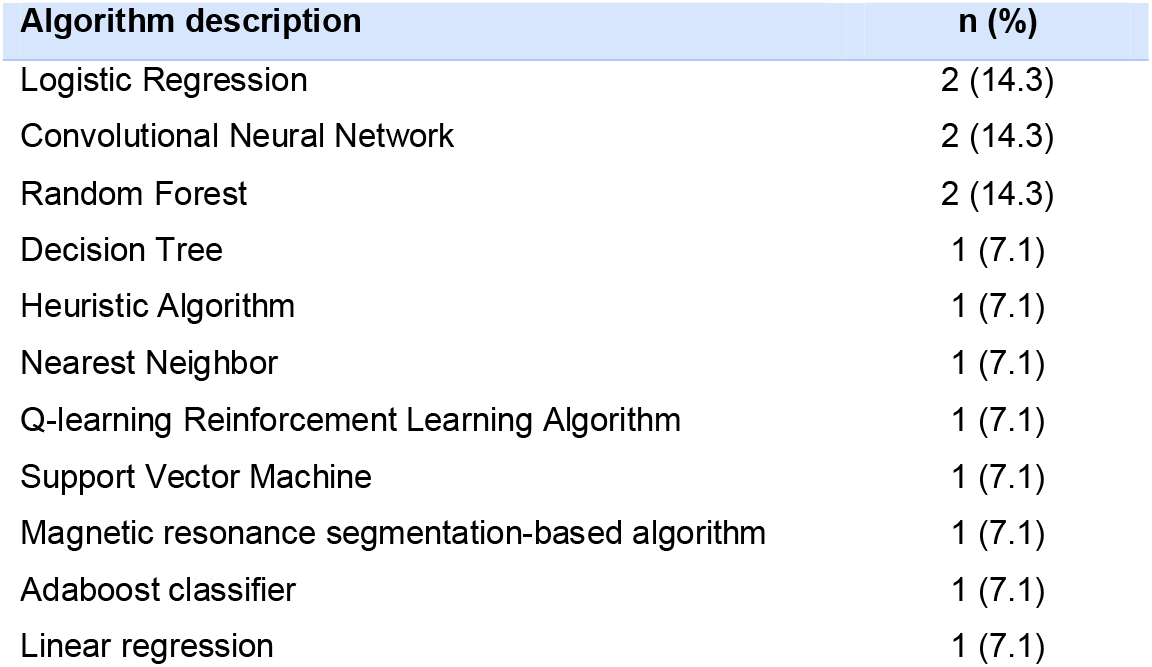
Prevalence of Machine Learning Algorithms in Neurosurgical Preoperative Planning.

**Figure 2.**
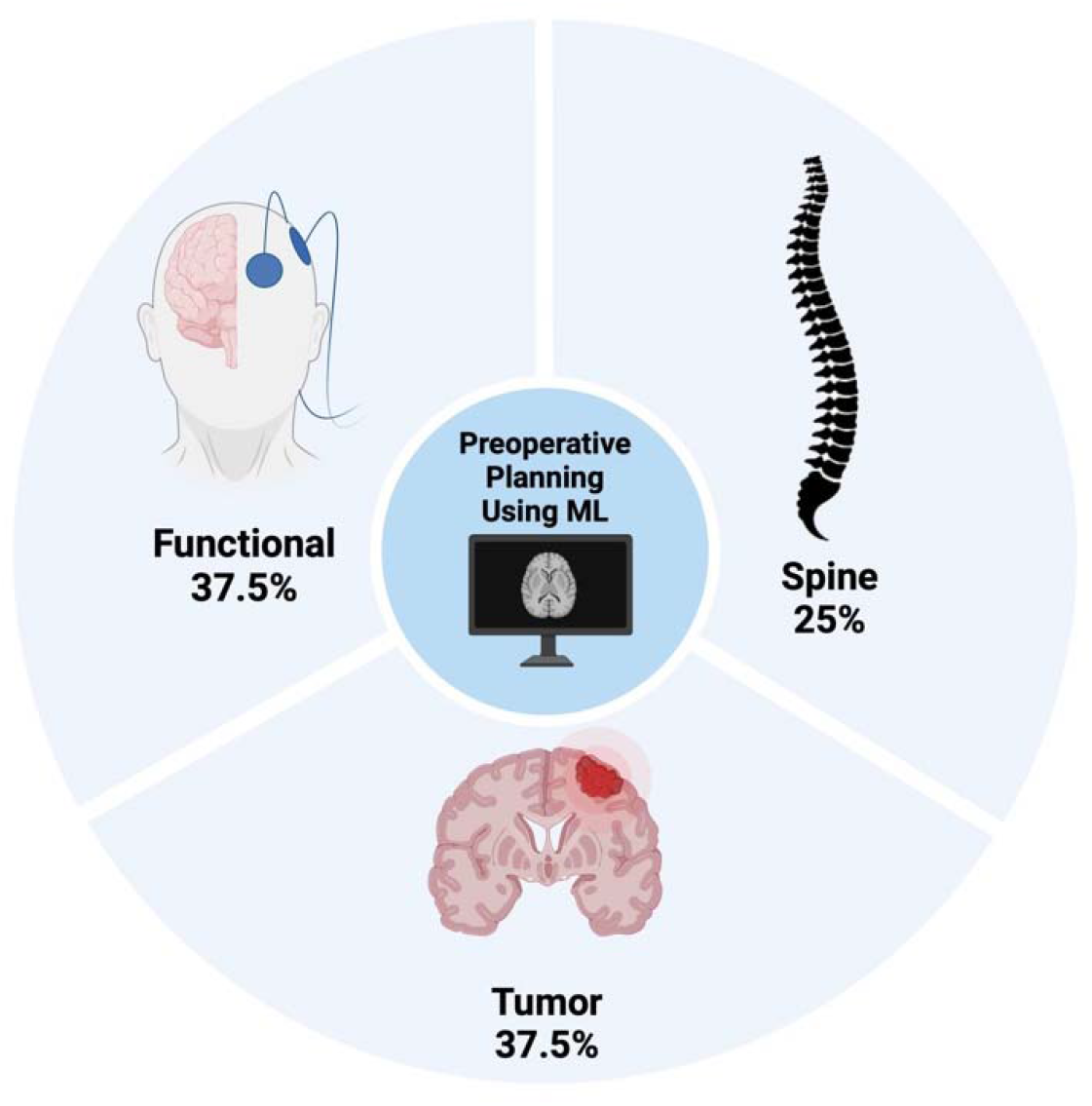
Distribution of Machine Learning Algorithms across Neurosurgical Specialties

**Figure 3.**
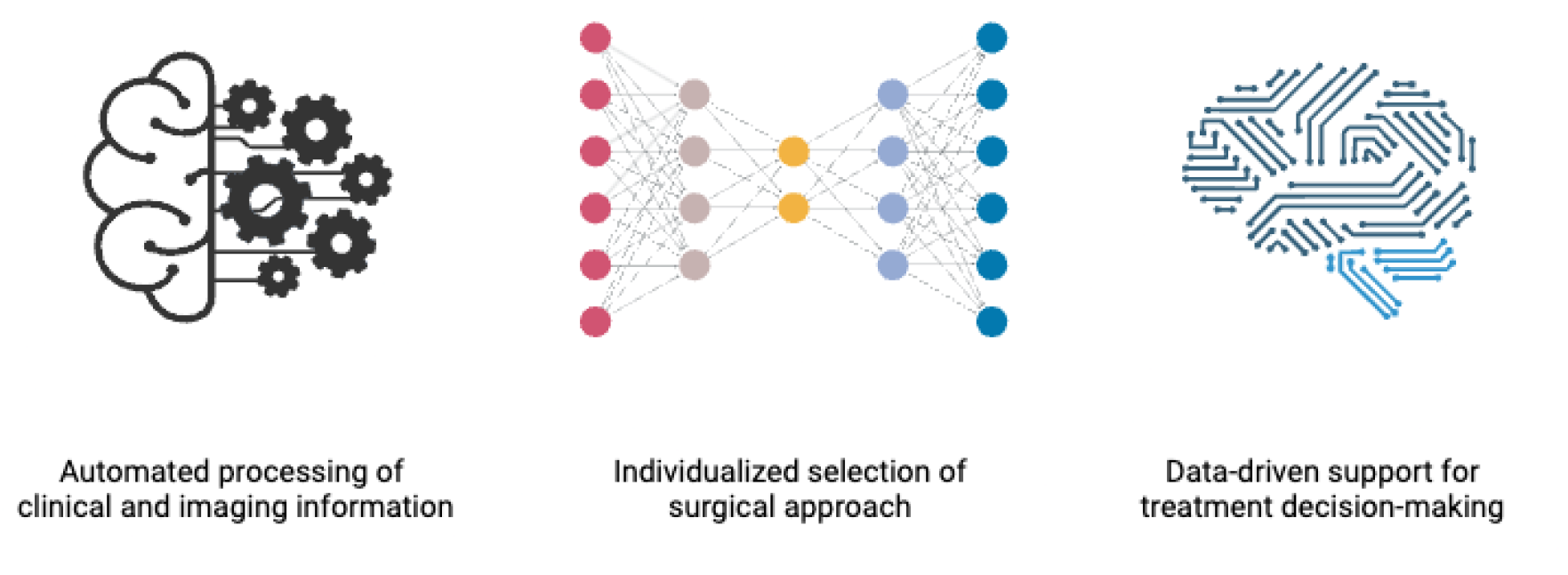
Main advantages of using Machine Learning during preoperative neurosurgical planning

## Discussion

In this study, the dynamic integration of ML is explored in neurosurgical planning, which is a field critical for enhancing patient safety and surgical precision. As dissected the use of various ML algorithms across eight studies involving 518 patients, it becomes evident that ML brings forth substantial advantages alongside significant challenges.

### Advantages and Prospects of ML Algorithms in Neurosurgical Planning

The profound impact of ML in neurosurgery lies in its ability to transform preoperative planning through a host of advantages, which this review details meticulously. Primarily, ML facilitates highly personalized surgical strategies. Algorithms such as convolutional neural networks, logistic regression, and random forests have the capability to process complex datasets and tailor surgical approaches to the specific anatomical and pathological characteristics of individual patients.^24^ This level of customization is paramount in neurosurgery, where the accuracy of the surgical approach can drastically affect patient outcomes.

Furthermore, ML significantly automates the processing of clinical data. This automation is a critical advancement as it alleviates the manual burden on neurosurgeons by quickly and accurately sifting through large volumes of patient data to extract relevant insights.^25^ Such efficiency not only speeds up the preoperative planning process but also enhances the reliability of the outcomes, reducing the likelihood of errors that might occur with manual data handling.^8,25^

Additionally, ML algorithms excel in supporting complex decision-making processes.^8,25^ They provide neurosurgeons with enhanced analytical capabilities, presenting data-derived insights and predictive analytics that help in making more informed, strategic decisions about surgical approaches and interventions.^8,25^ For instance, algorithms can predict the risk of specific preventable complications based on patient data, thereby aiding surgeons in choosing the safest and most effective surgical paths.^26–28^

ML also plays a pivotal role in risk assessment.^24,27^ By identifying potential surgical complications before they occur, ML algorithms contribute to a proactive approach to surgical planning.^26,28^ This predictive capability is invaluable in neurosurgery, where preemptive identification of risk factors can lead to significant improvements in surgical safety and patient outcomes.^29,30^

However, despite these substantial advantages, the implementation of ML in neurosurgery is not without its challenges.^25,27^ The success of ML algorithms depends heavily on the availability of extensive, high-quality data sets, which are often difficult to gather, particularly for less common conditions.^25,27^ Issues such as algorithmic bias and the challenge of generalizing results across diverse patient populations further complicate the effectiveness of ML applications.^8,28,29^ Moreover, the technical demands and resource intensity required to implement and maintain these systems pose significant hurdles, especially in settings with limited technological infrastructure.^24,27^

In addition, the complexity of machine learning models can make them less accessible to clinicians without specialized training in data science, potentially hindering broader adoption and trust in these tools.^8^ Ethical considerations, such as patient consent and data privacy, along with the evolving regulatory environment for AI technologies, underscore the need for careful management of these innovative tools within clinical practice.^25^

A notable example is the study by Baxter et al.,^25^ which utilized a CNN in a two-stage separable learning workflow for subthalamic nucleus segmentation. The initial phase employed a multi-resolution CNN to estimate the nucleus’s location, guiding the cropping of images to a smaller region for precise segmentation using a U-Net style architecture. This method outperformed traditional registration-based methods and included a human-computer interaction mechanism that significantly improved error correction by users. Therefore, the Baxter study illustrates the potential of ML to personalize surgical planning, enhancing surgical accuracy, minimizing risks, and tailoring interventions to specific anatomical details. Despite its success, this study also highlights the need for ongoing interaction and updates to ensure optimal algorithm performance. This necessity emphasizes the broader requirement for continuous maintenance, regular updates, and a collaborative approach involving clinicians and data scientists to ensure the efficacy and safety of these technologies in real-world settings.

### Recommendations and Analysis of Good Practices for Implementing ML Algorithms

It is important to note that regardless of the type of ML algorithm, authors are required to spend significant time validating and selecting the data for which the models are going to be trained. This implies that the retrieved information is representative of common clinical scenarios and not a pool of outliers. Moreover, it is advisable to leave a record of the authors who conducted these processes.^31^

In the case of poor-quality data (e.g., computed tomography scans with low image resolution), efforts must be made to optimize this crucial component, which can translate into a model yielding good performance.^32^

⍰

Before an algorithm is applied, data processing, training, and evaluation must be documented, replicated, and approved by specialists. This allows individuals interested in the ML algorithm to conduct audits and ensure that the model is accurate rather than a result of chance. Additionally, it enhances transparency in the process to ascertain whether best practices were used and if a thorough analysis was performed to eliminate overfitting.^31^

Once the models have been trained, their predictions need to be evaluated to understand the situations in which they are most and least successful. This process should also be carried out using new data before retraining the model. Depending on the specific scenario, a policy needs to be established for retraining the model when its performance deteriorates.

Special care should be taken when using tree-type models – as in the case of Gadot et al.^26^ – as they may have good performance but are prone to overfitting. In such scenarios, it is advisable to use ensemble algorithms, such as random forest, LightGBM, or XGBoost.^33^

It is important that the person in charge of training the models defines and generates the interpretation mechanisms (preferably visually). For instance, in a decision tree algorithm, trees can be graphed to understand its decision-making process.^34^ For convolutional neural networks, show heat maps in each prediction to understand which regions the algorithm focused on.^35^ In linear or logistic regression algorithms, evaluate the coefficients and intercept. In the “K Nearest Neighbor” algorithm, show which are the neighbors on which the algorithm bases its prediction.^36^ For more complex algorithms, the use of SHapley Additive exPlanations (SHAP) can be recommended to explain the result of the algorithm and even to evaluate how the features influence the model predictions.^37^

Finally, it’s crucial to ensure that the features used in training the models are ethically balanced to avoid introducing biases, such as in gender or age groups. This is important because certain medical conditions may have a higher correlation or prevalence in specific demographic cohorts.

In summary, while machine learning offers transformative potential for neurosurgical preoperative planning, realizing this potential requires navigating a landscape filled with both technological promise and significant challenges. Enhancing the robustness, transparency, and accessibility of ML applications will be crucial for their successful integration into neurosurgical practice, ensuring they improve patient care while adhering to the highest standards of ethical medical practice.

## Conclusion

ML algorithms for preoperative neurosurgical planning are being developed for efficient, automated, and safe treatment decision-making. Enhancing the robustness, transparency, and understanding of ML applications will be crucial for their successful integration into neurosurgical practice.

## Data Availability

Not applicable

