## Supplementary material for "Machine Learning Algorithms for Neurosurgical Preoperative Planning: A Comprehensive Scoping Review of the Literature": Tables

**Table 1. Characteristics of included studies, demographics, and clinical information.** AI: artificial intelligence; AIS: adolescent idiopathic scoliosis; CT: computed tomography; DBS: deep brain stimulation; DL: deep learning; ML: machine learning; MRI: magnetic resonance imaging; PC: principal components; VS: vestibular schwannoma; 3D: Tridimensional.

| **Author, Year** | **Country** | **Study design** | **Neurosurgical specialty** | **Patients (n)** | **Male/ Female** (n) | **Age (mean) (years)** | **Neurological condition of study** | **ML subset** | **Aim of using ML preoperatively** |
| --- | --- | --- | --- | --- | --- | --- | --- | --- | --- |
| Berlin, 2023^24^ | Germany | Retrospective | Spine | 100 | 20/80 | 14.6 | AIS | DL | Skeleton measurements (e.g., Cobb angle, coronal balance) relevant for AIS planning |
| Baxter, 2022^25^ | France | Experimental | Functional | 17 | 10/7 | NR | Neurological and neurodegenerative disorders | DL | Estimation of subthalamic nucleus localization for DBS stimulation |
| Dundar, 2022^8^ | Turkey | Retrospective | Tumor | 1 | NR | NR | Intracranial tumors | ML | Select cranial surgery approaches based on individual anatomic features |
| Gadot, 2022^26^ | USA | Retrospective | Tumor | 124 | 81/43 | 52.4 | Vestibular schwannomas | ML | Decision-making for active treatment vs MRI surveillance for patients with vestibular schwannomas |
| Kamer, 2021^27^ | Germany | Retrospective | Spine | 20 | 2/18 | 78.65 | Non‐or minimally‐ displaced fragility fractures of the sacrum | AI and CT based 3D statistical modeling | Planning of transsacral implant position |
| Coenen, 2019^28^ | Germany | Prospective | Functional | 8 | 5/3 | NR | Psychiatric disorders, specifically major depressive disorder and obsessive-compulsive disorder | ML | White matter tract tracking for DBS planning. |
| Gazit, 2016^29^ | Israel | Prospective | Functional | 76 | 37/39 | NR | Epilepsy | ML | Preoperative language area mapping |
| Zhai, 2021^30^ | China | Retrospective | Tumor | 172 | 37/135 | 52.8 | Meningioma | ML | Meningioma tumor consistency assessment prior to surgery |

**Table 2. Details related to ML algorithms, advantages and limitations.** AI: artificial intelligence; CNN: convoluted neural networks; CT: computed tomography; DBS: deep brain stimulation; DL: deep learning; dMRI: Diffusion magnetic resonance imaging; ML: machine learning; HAMLET: Hierarchical Harmonic Filters for Learning Tracts from Diffusion MRI; MRI: magnetic resonance imaging; STN: subthalamic nucleus; slMFB: superolateral medial forebrain bundle

| **Author** | **Algorithm Type** | **Preoperative ML algorithm description** | **Advantages of using ML preoperatively** | **Limitations of ML algorithms** |
| --- | --- | --- | --- | --- |
| Berlin, 2023^24^ | CNN | Automated algorithm containing a DL-CNN to at first identify different anatomical structures in anteroposterior spine X-rays and subsequently compute parameters based on the network’s output. | The reliability and speed offered by the AI-algorithm could contribute to the efficient analysis of large datasets (e.g., registry studies) and measurements in clinical practice. | The AI algorithm measured exclusively coronal parameters. |
| Baxter, 2022^25^ | CNN | Two-stage separable learning workflow for STN segmentation. The first part used a multiresolution CNN to determine an estimate of the subthalamic nucleus location, which was then used to heavily crop the input images to the much smaller region of interest. The second network then directly segmented these images using a U-Net style architecture. | Two-step segmentation significantly outperformed the comparative registration-based method currently used in clinics and approaches the fundamental limit on variability due to the image resolution. The human-computer interaction experiment showed that the additional interaction mechanism allowed by separating STN segmentation into two steps significantly improves the users' ability to correct errors and further improves performance. | The study coupled interaction for left and right STNs, which affected the timing and ease-of-use results. |
| Dundar, 2022^8^ | Heuristic algorithm and Q-learning reinforcement learning algorithm | In the first stage, a heuristic algorithm was used with MRI data to compute optimal surgical paths, avoiding critical structures in the brain and selecting the best ones based on reward and penalties.  In the second stage, the Q-learning reinforcement learning algorithm was employed with optimal linear paths as entry points. This algorithm searched for the best nonlinear routes, minimizing the risk of damage to critical brain structures | Personalized surgical planning with optimized, precise and accurate trajectories. | Manual segmentation and specification of anatomical points, difficulties with image fusion, lengthy processing time, and limited automatic segmentation. |
| Gadot, 2022^26^ | Decision tree and Random Forest | Decision trees were trained to predict the decision of active treatment or surveillance based on preoperative variables. A random forest with 500 trees was used to predict the decision of active treatment versus surveillance in patients with vestibular schwannoma.  These algorithms were trained and validated with the goal of identifying which preoperative variables had the most weight in the intervention decision and could be used to guide future surgical decision making. | Accurate prediction of intervention, identification of predictive factors, personalized treatment, support in decision making and reduction of clinical variability. | Dependency on training data, interpretability, limited generalization, algorithmic bias, computational and resource requirements, and need for continuous update. |
| Kamer, 2021^27^ | Linear regression | Authors trained, validated, and tuned classification models for the binary existence of S1 corridors and regression models for the numeric PC scores and the S1 diameter as responses. Models were trained using supervised ML. Predictor variables were the features. Response variables were the binary existences of S1 corridors, the S1 corridor diameter, and the PC scores as mentioned above. | Improved and facilitated clinical evaluation, therapeutic decision‐making, and treatment planning with lower treatment risks in patients affected by fragility fractures of the sacrum. | CTs with low image resolution (CTs with 2 and 2.5 mm in the z‐axis (=patient axis) compromised data processing and analysis. |
| Coenen, 2019^28^ | Image segmentation-based algorithm | The HAMLET algorithm used harmonic filters and ML to identify and represent the slMFB beam on individual dMRI images, facilitating preoperative surgical planning for DBS. | Tha algorithm provided a more accurate and objective representation of the slMFB anatomy in each patient, and eliminated the subjectivity associated with manual and deterministic tractography methods. Thus, facilitating more refined surgical planning by identifying and excluding unwanted fibers, which may improve clinical outcomes for patients. | Training data dependence, interpretation of results, overfitting, need for updating and maintenance, risks of algorithmic bias, and computational and resource cost. |
| Gazit, 2016^29^ | Logistic regression | The probabilistic logistic regression algorithm was used to predict language lateralization in patients with epilepsy before brain surgery, using fMRI data and other clinical and neuropsychological measures. This approach provided an accurate, non-invasive assessment of language lateralization, which helped guide surgical planning and improve outcomes. | The algorithm can improve the accuracy of diagnoses, optimize treatment plans, prevent complications, and improve surgical outcomes, benefiting both patients and healthcare providers. | Data quality, overfitting, interpretability, data privacy and security, algorithmic bias, computational resources, domain specificity, ethical and regulatory concerns. |
| Zhai, 2021^30^ | Logistic regression, Random Forest, Nearest neighbor, Support Vector Machine, and Adaboost classifier | MRI and regions of interest were delineated in meningioma images. The, the algorithm were trained to obtain the predictive accuracy of each model in terms of under the curve (AUC), sensitivity, specificity, and accuracy on an independent test data set. | The use of algorithms offered several advantages such as individualized prediction, non-invasive, improved precision, and facilitated decision-making. | Interpretability, generalization, data collection, bias and equity, update and maintenance. |

| **Algorithm description** | **n (%)** |
| --- | --- |
| Logistic Regression | 2 (14.3) |
| Convolutional Neural Network | 2 (14.3) |
| Random Forest | 2 (14.3) |
| Decision Tree | 1 (7.1) |
| Heuristic Algorithm | 1 (7.1) |
| Nearest Neighbor | 1 (7.1) |
| Q-learning Reinforcement Learning Algorithm | 1 (7.1) |
| Support Vector Machine | 1 (7.1) |
| Magnetic resonance segmentation-based algorithm | 1 (7.1) |
| Adaboost classifier | 1 (7.1) |
| Linear regression | 1 (7.1) |

**Table 3. Prevalence of Machine Learning Algorithms in Neurosurgical Preoperative Planning**
